# 18F-AV1451 PET imaging and white matter changes in progressive supranuclear palsy

**DOI:** 10.1101/19000596

**Authors:** Nicolas Nicastro, Patricia Vazquez Rodriguez, Maura Malpetti, William Richard Bevan-Jones, P. Simon Jones, Luca Passamonti, Franklin I. Aigbirhio, John T. O’brien, James B. Rowe

## Abstract

**Introduction:** Progressive supranuclear palsy (PSP) is characterized by deposition of straight filament tau aggregates in the grey matter of deep nuclei and cerebellum. White matter changes are increasingly documented as a feature of degenerative parkinsonism. We therefore examined the relationship between tau pathology (assessed via ^18^F-AV1451 positron emission tomography) and white matter integrity (using diffusion tensor imaging, DTI) in PSP.

**Methods:** Twenty-three people with clinically probable PSP-Richardson’s syndrome (age 68.8 ± 5.8 years, 39% female) and 23 controls underwent structural 3T brain MRI including DTI. Twenty-one patients also underwent ^18^F-AV145 PET imaging. DTI group comparisons were performed using Fractional Anisotropy (FA), Mean Diffusivity (MD) and Radial Diffusivity (RD). Voxel-wise white matter integrity was correlated with ^18^F-AV1451 binding in typical subcortical PSP regions of interest (i.e. putamen, pallidum, thalamus and midbrain). DTI and ^18^F-AV1451 imaging measures were correlated with clinical impairment.

**Results:** Widespread DTI changes in PSP subjects relative to controls (family-wise error FWE p<0.01) were observed. In PSP, higher ^18^F-AV1451 binding correlated with reduced white matter integrity in the bilateral internal capsule, corona radiata, and superior longitudinal fasciculus (FWE p<0.05). Association between cognitive impairment (ACER score) and white matter deficits were found in the genu of corpus callosum and cingulum (p<0.005).

**Conclusion:** This cross-sectional study demonstrates an association between *in vivo* proxy measures of tau pathology and white matter degeneration in PSP. Longitudinal studies and more specific PET probes for non-Alzheimer tauopathies are warranted to assess the complex interplay between microstructural changes and protein deposition in PSP.

## INTRODUCTION

Progressive supranuclear palsy (PSP) encompasses a broad spectrum of motor, cognitive, and behavioural impairments. These include akinesia, postural instability with early falls, oculomotor deficits, fronto-executive dysfunction, and neuropsychiatric features such as apathy and impulsivity ^1, 2^. Neuropathological studies have shown that PSP is characterized by intra-neuronal and astrocytic aggregation of toxic microtubule-associated protein tau (of 4-repeat tau isoforms, with straight filaments) ^3, 4^. Using ^18^F-AV1451 positron emission tomography (PET) allows to quantify and localise *in vivo* tau burden. ^18^F-AV1451 binding is consistently increased in the basal ganglia, thalamus, and dentate cerebellum in patients with PSP relative to controls ^5-7^, mirroring the neuropathological distribution ^4^. Albeit ^18^F-AV1451 affinity for tau inclusions in non-AD tauopathies is less than in AD ^8-10^, and it is not specific to tau ^11^, the distribution of ^18^F-AV1451 is distinct in PSP ^6^. Therefore, where high clinic-pathological correlations indicate likely PSP tau pathology, ^18^F-AV1451 can be used to quantify it and compare to other disease processes such as white matter pathology.

The integrity of white matter can be assessed with diffusion tensor imaging (DTI), on the basis of the restriction of motion of water molecules’ motion by natural ‘barriers’ such as the axon proteins and myelin sheets. Using fractional anisotropy (FA), mean and radial diffusivity (MD and RD, respectively), abnormal white matter has been consistently observed for PSP in the corpus callosum, superior longitudinal fasciculus (SLF), as well as in the internal capsule and fornix ^12-15^.

Despite the separate insights from DTI and ^18^F-AV1451 PET, the relationship between tau burden and white matter degeneration in PSP has remained unknown. We therefore directly compared DTI changes and ^18^F-AV1451 binding to assess the voxel-wise and regional associations between white matter damage and tau pathology in PSP, focusing on subcortical nuclei. We assessed clinical correlations of both ^18^F-AV1451 binding and DTI metrics with motor and cognitive impairment. Considering the extensive white matter damage observed in PSP in previous studies, we predicted a profound loss of microstructural integrity related to higher ^18^F-AV1451 binding, and that the severity of these changes are related to motor and cognitive deficits.

## METHODS

### Participants

The present study is part of the Neuroimaging of Inflammation in MemoRy and Other Disorders (NIMROD) protocol (Bevan-Jones *et al*., 2017). We included 23 participants with probable PSP (PSP-Richardson syndrome) who were recruited according to NINDS-SPSP 1996 criteria ^3^ but reconfirmed as meeting the current clinical diagnostic criteria of probable PSP-RS ^2^. 23 similarly aged healthy controls were also recruited, with MMSE >26/30, absence of regular memory complaints, and no history of major neurological, psychiatric or significant medical illness. Patients were identified from the regional specialist PSP-clinic at the Cambridge University Hospitals NHS Trust. Healthy controls were recruited via the Dementias and Neurodegenerative Diseases Research Network volunteer register. Informed written consent was obtained in accordance with the Declaration of Helsinki. The study received a favourable opinion from the East of England Ethics Committee (Cambridge Central Research, Ref. 13/EE/0104). Clinical and cognitive assessment included MMSE and revised Addenbrookes Cognitive Examination (ACER), and the PSP rating scale (PSP-RS) for PSP subjects ^16^.

### MRI acquisition and preprocessing

Participants underwent MRI imaging acquired on a 3T scanner (Siemens Magnetom Tim Trio) using a magnetization-prepared rapid gradient echo (MPRAGE) T1-weighted sequence (repetition time = 2300 ms, echo time = 2.98 ms, field of view = 240 x 256 mm^2^, 176 slices, flip angle = 9°, isotropic 1mm voxels).

Diffusion-weighted images were acquired with a 65-direction encoding scheme, 2 mm thickness, TE = 106 ms, TR = 11700 ms, field of view = 192 x 192 mm^2^. 64 volumes were acquired with a b-value of 1000 s/mm^2^ following an initial volume with a b-value of 0 s/mm^2^. The data were preprocessed with the FSL 6.0 software package (http://www.fmrib.ox.ac.uk/fsl) using FSL Diffusion Toolbox. First, the series was adjusted for head movement and eddy currents using *eddy* and realigned to the b0 image. A brain mask was then produced by applying the *Brain Extraction Tool* (*BET*) to the (mean) b0 image. *DTIfit* was next used to independently fit the diffusion tensor at each voxel, resulting in the derivation of FA, MD and RD maps.

### PET acquisition and preprocessing

21 PSP participants underwent ^18^F-AV-1451 PET imaging. The radioligand was prepared at the Wolfson Brain Imaging Centre (WBIC), University of Cambridge. PET scanning was performed on a GE Advance PET scanner or a GE Discovery 690 PET/CT (General Electric Healthcare, Chicago, Illinois, USA). A 15-min ^68^Ge/^68^Ga transmission scan was used for attenuation correction. The emission protocol was as follows: 90 min dynamic imaging following a 370 MBq ^18^F-AV1451 injection. Each emission image series was aligned using SPM8 to correct for patient motion during data acquisition (www.fil.ion.ucl.ac.uk/spm/software/spm8). The mean aligned PET images were rigidly registered to the T1-weighted image to extract values from both the Hammers atlas regions of interest (ROIs) and those in a reference tissue defined in the superior grey matter of the cerebellum using a 90% grey matter threshold on the grey matter probability map produced by SPM8 smoothed to PET resolution ^17^. Regional PET data were corrected for CSF partial volumes. ^18^F-AV1451 non-displaceable binding potential (BP_ND_) was determined for each region of interest using a basis function implementation of the simplified reference tissue model (SRTM) ^18^.

### Statistical analyses

Demographic data were analyzed with Stata software Version 14.2 (College Station, TX). Assessment of distribution for continuous variables was performed with Shapiro–Wilk test and visualization of histogram plots, followed by *t* test or Mann–Whitney U, accordingly. Categorical variables were compared with Chi-Square test. Statistical significance was considered when p<0.05.

The DTI pipeline aligned each subject’s FA image to a pre-identified target FA image (FMRIB_58) ^19^. Affine registration into the Montreal Neurological Institute (MNI) MNI152 space was performed. A mean FA image and skeleton were created from all subjects and each individual’s FA image was then projected onto the skeleton, with a threshold of 0.2 applied to the mean skeleton in order to include white matter tracts that were common across all subjects and to exclude voxels that may contain grey matter or CSF. The aligned DTI parameter map of each subject was then back-projected onto the mean skeleton. In addition, other DTI parameters (i.e., MD and RD) were aligned by applying the original FA non-linear spatial transformations to the corresponding datasets and projecting them onto the mean FA skeleton. Subsequently, the *randomise* function in FSL was implemented to identify group differences in DTI metrics between PSP subjects and controls, using an independent sample t-test, with age and gender as covariates of no interest.

Similarly, voxel-wise correlational models between DTI measures and average (of left and right) ^18^F-AV1451 binding in specific ROIS in PSP subjects included age, gender, and disease severity (as measured with PSP-RS) as covariates of no interest. Subcortical ROIs’ ^18^F-AV1451 BP_ND_ significantly increased in a previous PSP study were treated as the independent variable for each DTI parameter. Statistical significance was determined using non-parametric permutation testing (n=10’000 permutations), applying threshold-free cluster enhancement (TFCE) and adjusting for multiple comparisons with family-wise error (FWE) p< 0.05 ^20^. Anatomical labelling of significant TBSS white matter clusters were facilitated using the John Hopkins – ICBM white matter atlas, available as part of the FSL package.

For PSP subjects, correlation analyses between clinical scales (ACER and PSP-RS) and regional ^18^F-AV1451 BP_ND_ or DTI metrics were performed, using multiple linear regressions correcting for age and gender. For DTI, extraction of the mean FA/MD/RD values in the white matter tracts that showed significant deficits in PSP participants compared to controls was performed. Bonferroni-corrected p-value (p = 0.05/n with n being the number of distinct white matter tracts tested) was considered as significant for clinical measures and DTI correlations.

## RESULTS

### Demographics

Demographic and clinical characteristics of PSP and controls participants are shown in **Table 1**. Both groups were comparable in terms of age, gender and education attainment. As expected, cognitive scores (i.e., MMSE and ACER) were lower in the PSP group (p=0.01 and p<0.001, respectively).

**TABLE 1.**

|  | **PSP (n=23)** | **Controls (n=23)** | **p-value** |
| --- | --- | --- | --- |
| **Age, years** | 68.8 ± 5.8<br>(52-79) | 68.7 ± 7.3<br>(55-81) | ns * |
| **Female participants** | 39% (9/23) | 48% (11/23) | ns § |
| **Education, years** | 12.2 ± 2.1<br>(10-17) | 13.5 ± 2.5<br>(9-17) | ns * |
| **MMSE** | 26.0 ± 4.5<br>(13-30) | 28.7 ± 1.1<br>(26-30) | 0.01 # |
| **ACER** | 78.2 ± 16.5<br>(36-95) | 91.3 ± 6.3<br>(75-99) | < 0.001 # |
| **PSP-RS** | 48.0 ± 16.3<br>(25-77) | -- | na |

Clinical characteristics of included subjects. Values are mean ± standard deviation (range).
Student t-test \*, Chi-Square test §, Mann-Whitney U test #. ns = not significant p>0.05
uncorrected, na = not applicable

### DTI group comparisons

Voxel-based TBSS analyses confirmed that relative to controls, PSP subjects had diffuse white matter damage in the corpus callosum (genu, body, and splenium), bilateral internal capsule (anterior and posterior limb), corona radiata (anterior, superior, and posterior parts), posterior thalamic radiations, cingulate white matter, SLF, sagittal stratum, uncinate fasciculus, and fornix (cres)/stria terminalis (TFCE p< 0.01). These abnormalities were consistent across the three DTI metrics (i.e., decreased FA, increased MD and RD in PSP) (**Figure 1**).

**FIGURE 1.**
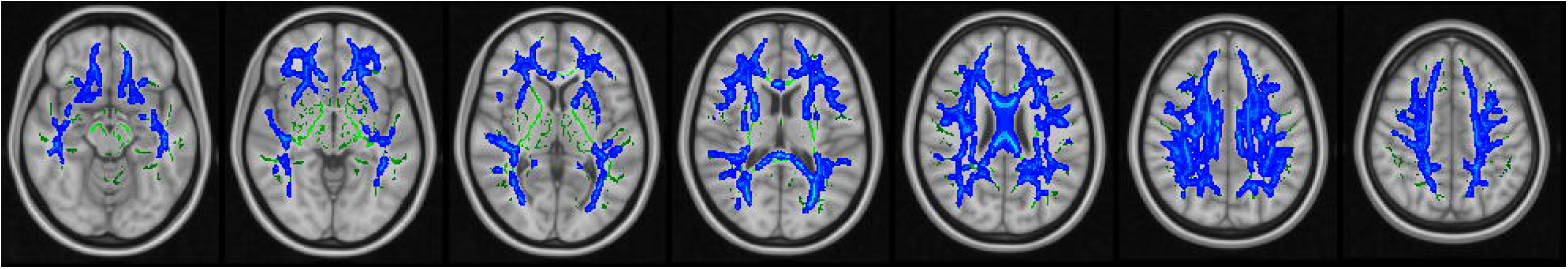
DTI TBSS group comparisons showing increased radial diffusivity (blue) in PSP subjects compared to Control participants overlaid on the common white matter skeleton (green). Results are family-wise error corrected p < 0.01.

### Voxel-wise correlation of DTI imaging and ^18^F-AV1451 binding

As shown in a previous publication ^6^, subcortical ROIs for which PSP subjects had significantly higher ^18^F-AV1451 BP_ND_ relative to controls included putamen, thalamus, pallidum and midbrain (all p<0.02). We therefore used individual ROI ^18^F-AV1451 BP_ND_ values to assess their voxel-wise correlation with DTI imaging. For each ROI, higher ^18^F-AV1451 BP_ND_ was correlated with white matter measures (e.g., increased MD/RD or decreased FA) in the retro-lenticular part of internal capsule, superior and posterior corona radiata, posterior thalamic radiations and SLF (TFCE p<0.05) (**Figure 2**). Higher ^18^F-AV1451 binding in the thalamus and midbrain was associated with additional increased MD/RD and reduced FA in the body of corpus callosum, anterior corona radiata, cingulate white matter, fornix (cres) / stria terminalis and superior fronto-occipital fasciculus (TFCE p<0.05).

**FIGURE 2.**
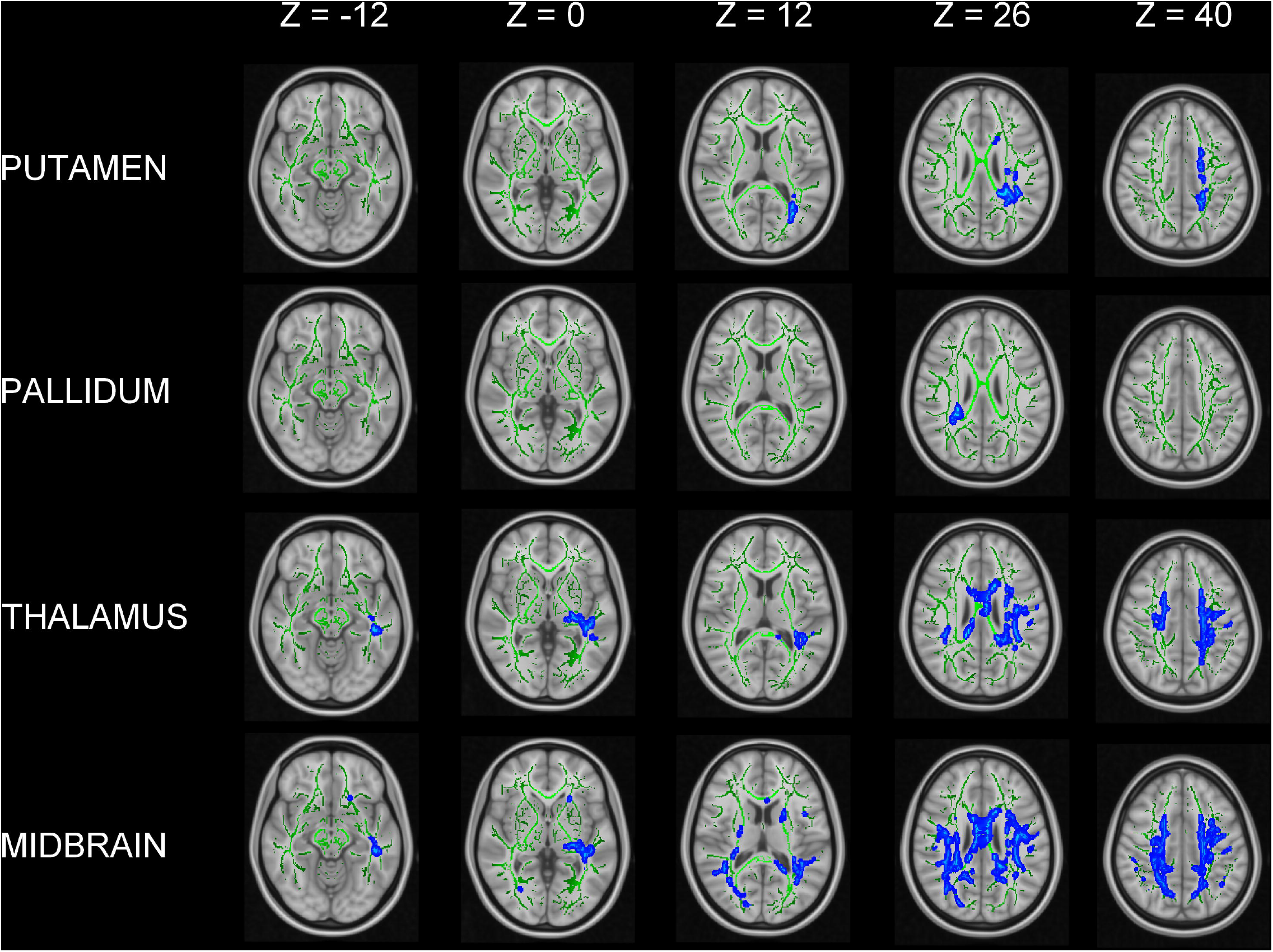
TBSS voxel-wise positive correlation of DTI impairment (increased RD) in association with higher regional ^18^F-AV1451 BP_ND_ in putamen, pallidum, thalamus and midbrain for PSP subjects. MNI Z-axis coordinates are shown. Results are FWE-corrected p < 0.05.

### Clinical correlations with ^18^F-AV1451 and DTI metrics

We did not find significant univariate correlations between regional ^18^F-AV1451 BP_ND_ and either ACER or PSP-RS (all p>0.2). Lower ACER score was correlated with white-matter deficits in the genu of corpus callosum (FA: t = 3.85, p = 0.001; RD: t = −3.45, p = 0.003) and cingulate white matter (FA: t = 3.96, p = 0.001). We did not find significant correlations between DTI metrics and PSP-RS (all p > 0.015).

## DISCUSSION

In this study, we demonstrate that higher ^18^F-AV1451 binding in subcortical regions affected by PSP was correlated with changes in the diffusion tensor characteristics of white matter. We interpret these correlations in terms of a correlation between tau pathology and microstructural integrity of white matter, while recognizing important caveats below.

In comparing PSP patients with controls independently of ^18^F-AV1451 binding, extensive white matter impairment was found, including the corpus callosum, internal capsule, corona radiata, posterior thalamic radiations, SLF, and fornix. These results are in keeping with previous observations ^12, 14, 15^, which suggest a key role of white matter changes in the pathophysiology of PSP, especially the PSP-Richardson syndrome ^21^.

We also observed that ^18^F-AV1451 in high-binding regions for PSP (i.e. thalamus, midbrain, putamen and pallidum) was positively correlated with increased MD/RD and reduced FA in motor tracts (corona radiata, internal capsule), posterior thalamic radiations and SLF, the latter being particularly involved in spatial attention, oculomotor function and motor behaviour ^22^. In relation to higher ^18^F-AV1451 binding in the thalamus and midbrain, we observed changes in additional white matter regions. These included the body of corpus callosum, whose fibres pass through the corona radiata to reach the brain surface, the cingulum (which is densely connected to the thalamus and spinothalamic tract) and superior fronto-occipital fasciculus. These cross-modal association shed light onto the intricate *in vivo* relationship between the integrity of connecting fibres and ^18^F-AV1451 binding. MD and RD seem able to detect more subtle changes than FA in PSP ^23-25^. However, it is still unclear whether microstructural damage is a direct consequence of tau pathology in the white matter (as observed *post mortem*) or consecutive to Wallerian degeneration.

We did not find a correlation between motor (i.e., PSP-RS) or cognitive (i.e., ACE-R) severity and regional ^18^F-AV1451 binding. This is in accordance with Schonhaut et al. ^7^ but contrasts with Smith et al., who found a correlation between pallidal ^18^F-AV1451 binding and PSP-RS^9^. One explanation could be the relatively small sample sizes or early disease stage of our patients. In AD, ^18^F-AV1451 binding in the temporal cortex was negatively related to temporal (especially its medial portion) cortical thickness change ^26^. In addition, Bejanin *et al*. showed that tau deposition was related in a region-specific manner to cognitive decline ^27^. That ^18^F-AV1451 is a more sensitive marker of paired helical filaments than straight filamentous might also play a role.

We did find that lower cognitive performance was significantly associated with impaired diffusion metrics in the genu of corpus callosum and cingulum. These highlight the prominent role of frontal dysfunction in PSP ^28, 29^ as both the genu of corpus callosum and cingulum are densely connected to the frontal lobe ^30^. In a previous study, Piattella *et al*. showed that MMSE score in PSP was correlated with whole white matter mean FA ^31^.

Our study is not without limitations. First, our results would require confirmation in larger and independent samples. Second, the present relationship between ^18^F-AV1451 and white matter integrity is based on a cross-sectional design: longitudinal studies are required to fully assess the spatial and temporal interplay between white matter integrity and ^18^F-AV1451 in PSP, and test causal models of pathology in humans. In addition, ^18^F-AV1451 off-target binding is a caveat to our interpretation. Neuromelanin-containing cells in the substantia nigra and monoamine oxidase in the striatum are bound by this ligand ^32^. However, we recently showed that off-target binding in basal ganglia, cortex or adjacent white matter is not sufficient an explanation, as *post mortem* data did not show relevant neuromelanin-containing cells ^6^. The very high clinic-pathological correlations in PSP-Richardson syndrome preclude TDP43 ^11^ or concurrent AD-pathology as an alternative explanation, noting that the six patients in our study who have come to post mortem examination were confirmed as PSP without significant dual pathology.

In conclusion, we present evidence of the *in vivo* association between white matter integrity and ^18^F-AV1451 in PSP. Longitudinal studies would be helpful to determine the dynamic changes occurring for both tau deposition and microstructural integrity. Tau PET probes more specific and sensitive for straight filaments would also enable confirmation of the complex interplay between cortical tau aggregation, white matter tracts degeneration and disease progression in PSP.

## Data Availability

data is available to researchers upon request

## APPENDIX 1 AUTHORS CONTRIBUTIONS

APPENDIX 1 – AUTHORS CONTRIBUTIONS
| Authors | Location | Roles and Contributions |
| --- | --- | --- |
| Nicolas NICASTRO | University of Cambridge | Data analysis and interpretation, draft and revision of the manuscript |
| Patricia VAZQUEZ<br>RODRIGUEZ | University of Cambridge | Data collection, critically revised the manuscript for important intellectual content |
| Maura MALPETTI | University of Cambridge | Data analysis, revision of the manuscript |
| William Richard<br>BEVAN-JONES | University of Cambridge | Data collection and analysis, revision of the manuscript |
| P. Simon JONES | University of Cambridge | Data collection and analysis, revision of the manuscript |
| Luca PASSAMONTI | University of Cambridge | Data collection and analysis, revision of the manuscript |
| Franklin I.<br>AIGBIRHIO | University of Cambridge | Study design, revision of the manuscript |
| John T. O'BRIEN | University of Cambridge | Study design and concept, data interpretation, revision of the manuscript |
| James B. ROWE | University of Cambridge | Study design and concept, data interpretation, revision of the manuscript |

## ACKNOWLEDGEMENTS

Thanks to our volunteers for participating in this study and to the radiographers at the Wolfson Brain Imaging Centre and PET/CT Unit, Addenbrooke’s Hospital, for their invaluable support in data acquisition. We thank the NIHR Dementias and Neurodegenerative Diseases Research Network for help with subject recruitment. We also thank Dr Istvan Boros, Joong-Hyun Chun, and WBIC RPU for the manufacture of 18F-AV-1451. We thank Avid (Lilly) for supplying the precursor for the manufacturing of AV-1451 for use in this study.

## ETHICAL APPROVAL

The present study was performed in agreement with the Declaration of Helsinki and its further amendments. Approval was obtained from Ethics Committee from East of England (Cambridge Central Research, Ref. 13/EE/0104).

## INFORMED CONSENT

Informed consent has been obtained from all participants in the present study.

